# Germline Variant Profiling of NA12878 Chromosome 22 Identifies the Loss-of-Function CYP2D6*3 and *4 Pharmacogenetic Alleles

**DOI:** 10.64898/2026.11.19.26368475

**Authors:** Suvajit Mallick, Bhupender Singh

## Abstract

The approach used in this project to call germline variants from short-read sequencing data forms the basis of a great deal of current human genetics, whether it be in population studies or in diagnostic testing. A full germline variant-calling workflow according to the GATK Best Practices was constructed and carried out using public whole-genome sequencing data from the reference sample NA12878. In order to comply with the limitations of a laptop (an Intel Core i3, 8 GB of RAM, Ubuntu via WSL2), the analysis was limited to chromosome 22. The reads for chromosome 22 were streamed directly from the 1000 Genomes high-coverage CRAM file (run ERR3239334), converted into paired FASTQ files, checked using FastQC and MultiQC, and then cleaned with fastp. The cleaned reads were aligned to a GRCh38 reference containing only chromosome 22 using BWA-MEM, after which they were sorted, duplicates were marked and the base quality scores were recalibrated before variants were called with GATK HaplotypeCaller. The GATK-recommended hard filters were applied separately to SNPs and indels, and the variants that passed these filters were annotated using the Ensembl Variant Effect Predictor (VEP) and summarised with a Python script.

From the 81,599 raw calls, 73,570 (90.2%) survived the filtering process, and the transition/transversion ratio for the SNPs that passed filtering was 2.04. Of the annotated variant alleles, 98.9% were already present in the catalogue, 734 affected the coding sequence in 270 genes, and no variant corresponded to a pathogenic entry in ClinVar. Examination using IGV verified the read support for the selected high-impact calls, including the variants that define the CYP2D6*3 and *4 alleles, consistent with the published genotype of NA12878. The main limitations were a reduced effective depth of 8.6x, resulting from an incomplete read-collation step which retained about 31% of the read pairs, and the use of a chromosome-only reference.

## Introduction

### Next-generation sequencing and variant calling

It is now routine, thanks to next-generation sequencing (NGS), to sequence an entire human genome in just a few days at a cost which twenty years ago would have been unimaginable [1]. Sequencing platforms of the type Illumina NovaSeq generate hundreds of millions of reads, each of which is only 100 to 150 bases long, and these individual reads by themselves are not significant. They must be examined, aligned to a reference genome and then compared with it before any biological questions can be addressed. The process of identifying the positions at which a sample differs from the reference is known as variant calling, and it is at the heart of most human genetics research, whether it is involved in population studies or in diagnostic testing [2].

Variant calling is not carried out by a single program but involves a series of steps, and an error at any one of them is passed on to the next stage. Bases of low quality may appear to be mutations, duplicated reads can increase the evidence for a false variant call, and reads that have been placed in the incorrect section of the genome can result in the detection of variants that are not actually there. In response to these issues, the Genome Analysis Toolkit (GATK), developed at the Broad Institute, provided a range of tools together with a recommended workflow, commonly referred to as the GATK Best Practices, which has since become a standard reference for germline analysis [3–5].

The project is concerned with germline variants, which are the inherited differences found in all the cells of an individual. Since humans have two copies of each autosome, a true germline variant is expected in about half of the reads that include it (in the case of heterozygosity) or in almost all of them (when homozygous). This simple expectation proved to be a useful way of checking the results at every stage of the work.

### The reference sample NA12878

NA12878 is a woman with northern European ancestry from a family in Utah (belonging to the CEU population) whose cell line has been investigated by both the HapMap and the 1000 Genomes projects. Her DNA has been sequenced on almost all of the major platforms, and the Genome in a Bottle consortium used her genome to create one of the earliest high-confidence benchmark collections of SNPs and indels [6]. She took part in the 1000 Genomes Project [7], and her genome was subsequently resequenced to an average coverage of about 30x by the New York Genome Center as part of the expanded high-coverage 1000 Genomes dataset [8]. There is a practical benefit when working with a genome that has been so extensively studied: it is possible to compare some of the results, for example the overall variant profile or her pharmacogenetic genotypes, with previously published findings.

### Why chromosome 22

The human genome consists of approximately 3.1 billion bases, and the amount of memory and storage required to process a 30x whole genome exceeds that which the laptop used in this case could offer. It was for this reason that the analysis was restricted to chromosome 22. In the GRCh38 assembly [9], this chromosome has a length of about 50.8 Mb, and a large part of its short arm is an unsequenced gap. Although the chromosome is small, chr22 is gene-dense and includes areas that are known to be problematic for short reads, such as the low-copy repeats in 22q11, the immunoglobulin lambda locus and the drug-metabolism gene CYP2D6 together with its neighbouring pseudogene CYP2D7. It thus provides both simple and difficult areas for a variant-calling pipeline.

### Sequencing data and file formats

The FASTQ record produced by Illumina instruments contains the base sequence and a Phred-scaled quality value for each base, defined by the equation Q = −10 log10(P) where P represents the probability that the base is incorrect; Q20 and Q30 thus relate to error rates of 1% and 0.1% respectively [1]. Once alignment has been carried out, the reads are stored in the Sequence Alignment/Map (SAM) format or in its compressed binary version, BAM. The CRAM format provides additional compression by recording only the differences between each read and the reference sequence, which is why a copy of the reference sequence is required in order to read a CRAM file. Samtools and BCFtools are the standard tools used for working with these alignment files and the variant call files (VCF/BCF) that are generated later [10].

### Read quality control and trimming

FastQC evaluates the quality of the raw reads using various modules, including per-base quality, GC content, duplication level and adapter content, and assigns to each module a pass, warning or failure flag [11]. MultiQC gathers the results from FastQC and a number of other tools into a single report, which makes it simple to compare different samples or to observe the effect of a processing step [12]. Trimming involves the removal of the low-quality ends of the reads and of any remaining adapter sequences. Trimmomatic carries out trimming using a sliding-window quality filter and includes an optional adapter-clipping step which requires a file containing the adapter sequences [13]. fastp combines quality filtering, adapter trimming and reporting into one fast program. With paired-end data it is able to detect adapters from the overlap between the two reads of a pair without being given the sequence, and it can also identify overrepresented sequences [14].

### Read alignment

An aligner must be able to quickly process hundreds of millions of reads while allowing for mismatches as well as small insertions or deletions. BWA makes use of the Burrows–Wheeler Transform and an FM-index of the reference sequence in order to enable fast searching with only moderate memory requirements [15]. The BWA-MEM algorithm, which is based on maximal exact matches, is intended for reads that are 70 bp or longer and is the aligner recommended in the GATK Best Practices [5,16]. A mapping quality (MAPQ) is assigned to each aligned read and this expresses the confidence level of the aligner regarding the read’s position; reads that come from repeated or duplicated regions are given a low MAPQ since they match just as well in more than one location. GATK also requires read-group tags (which indicate the sample, library, platform and unit) so that the reads from different runs can be treated separately [5].

### Post-alignment processing

The aligned reads are sorted according to their genomic coordinates and are indexed in such a way that various programs can go directly to any specific region; duplicate reads, which result from PCR amplification during library preparation or from optical effects on the flow cell, are identified by having the same starting position and are marked instead of being deleted, so that subsequent tools can ignore them while the file remains intact [4]. Base quality score recalibration (BQSR) then adjusts for the systematic errors in the quality values given by the sequencer by modelling the observed mismatch rate as a function of the reported quality, the position in the read (cycle) and the surrounding sequence context, while at the same time excluding sites of known variation so that true polymorphisms are not treated as errors [4,5].

### Variant discovery and filtering

Early variant callers looked at each position separately. The GATK HaplotypeCaller, on the other hand, first identifies active regions, then reassembles the reads in those regions into candidate haplotypes and employs a pair hidden Markov model to determine how well each read supports each haplotype. As a result of this local reassembly, indel calling in particular is more accurate [17]. Each call is recorded in a VCF file together with annotations that explain the evidence, including the quality by depth (QD), two measures of strand bias (FS and SOR), the root-mean-square mapping quality (MQ) and the rank-sum tests which compare the reads supporting the reference allele with those supporting the alternative allele.

Raw calls still give rise to false positives. The variant quality score recalibration (VQSR) develops a filtering model using training data, but in order to do so it requires a large number of variants, usually obtained from many samples or complete genomes. For a single sample or a small region, GATK suggests using hard filtering with fixed thresholds on the annotations, and this should be done separately for SNPs and indels [5]. A common way of checking the results is to look at the transition-to-transversion (Ti/Tv) ratio for the SNPs, which is approximately 2.0 to 2.1 for whole-genome data; a considerably lower value indicates that there are many false positives [4].

### Functional annotation and interpretation

Nothing much can be said about the biology based just on a variant’s position. The Ensembl Variant Effect Predictor (VEP) attaches each variant to the overlapping transcripts and designates Sequence Ontology consequence terms, for example missense, synonymous or intronic, along with an impact class which is classified as HIGH, MODERATE, LOW or MODIFIER [18]. It is important to take into account the population frequency, since a variant found in many healthy individuals is probably not going to cause a serious disease. The Genome Aggregation Database (gnomAD) gives allele frequencies from around 76,000 genomes [19]. AlphaMissense makes use of a deep-learning model to predict how likely it is that a missense change will be harmful [20], and ClinVar gathers the clinical interpretations submitted by testing laboratories [21]. The Integrative Genomics Viewer (IGV) and its web version igv.js are still the standard method of examining the reads associated with a call [22,23].

### Difficult regions and the CYP2D6 locus

Short reads have difficulty when the genome includes nearly identical copies of the same sequence. The gene CYP2D6, which is responsible for the metabolism of a large proportion of commonly used drugs, is located next to the pseudogene CYP2D7, with which it has about 94% sequence identity, and the region is frequently found to have deletions, duplications and also hybrid genes that combine CYP2D6 and CYP2D7 [24]. Because general-purpose genotyping tools are unable to resolve such structural variations, specialised tools like Cyrius have been developed in order to genotype CYP2D6 from whole-genome data [24]. The Genetic Testing Reference Material programme (GeT-RM) of the US Centers for Disease Control and Prevention issues consensus CYP2D6 genotypes for the Coriell reference samples, including NA12878 [25].

### Rationale for the present work

The pipelines that have been published generally assume that access to a computing cluster is available; for a person working on a personal computer, it is possible to carry out every step of the GATK workflow by limiting the analysis to a single chromosome of a well-characterised sample and at the same time be able to compare the results with existing published knowledge. The drawbacks of this method, particularly those associated with the use of a chromosome-only reference, are examined in the Limitations section.

### Scope of this study

The project was based on an eight-week plan which included data retrieval, quality control and trimming, alignment, BAM clean-up and validation, base quality score recalibration, variant discovery using hard filtering, functional annotation, and the preparation of this manuscript. This study outlines each of the steps taken, the reasons for the decisions made, the problems that occurred during the process and the way in which they were dealt with, as well as what the final set of variants shows.

### Research objectives

The project had the following objectives:

1. To set up a reproducible Conda-based environment for germline variant calling on a Windows laptop via WSL2.
2. To get the NA12878 sequencing reads for chromosome 22 in an efficient manner without having to download the whole genome.
3. To assess raw read quality and remove adapters, low-quality bases and overrepresented sequences.
4. To align the cleaned reads to the GRCh38 reference to generate a sorted, duplicate-marked and indexed BAM file.
5. To recalibrate the base quality scores and document the effect with before and after plots.
6. To call germline SNPs and indels using GATK HaplotypeCaller and apply the standard hard filters.
7. To annotate the passing variants with Ensembl VEP and then summarise them using a Python script.
8. To look at a selection of the variants in IGV and record the limitations of the approach.

## Methods

### Computing environment

All work was performed on a laptop (Intel Core i3-1115G4, 8 GB RAM) running Ubuntu via WSL2. Project files were stored on the D: drive (/mnt/d/NGS_Project/) as the system drive had very little free space. Software was installed from the Bioconda and conda-forge channels [26] into a Conda environment called variant-calling (Python 3.10). MultiQC was installed via pip as conda reported a dependency conflict and R was installed in a separate environment (r-plots) to avoid having to change the main environment. Table 1 lists the software versions and Figure 1 provides an overview of the workflow.

**Figure 1:**
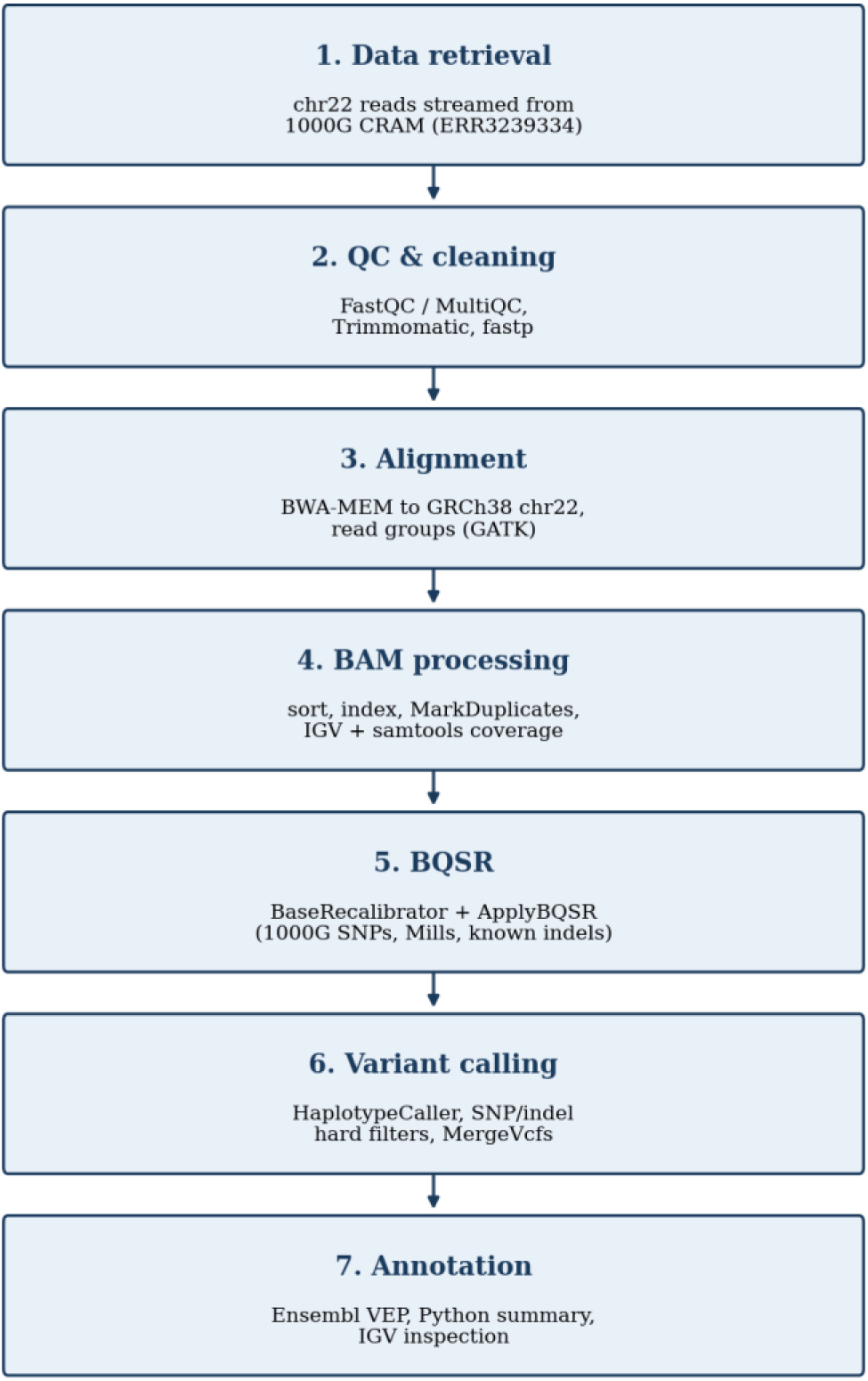
Overview of the variant-calling workflow followed in this project.

**Table 1:** Software used in this project.

| Tool | Version | Purpose |
|---|---|---|
| samtools | 1.23.1 | Read extraction, format conversion, sorting, indexing, statistics |
| bcftools | 1.24 | VCF subsetting, indexing, statistics, chromosome renaming |
| FastQC | 0.11.8 | Read quality reports |
| MultiQC | 1.35 | Combined quality-control report |
| Trimmomatic | 0.40 | Quality trimming (first pass) |
| fastp | 1.3.6 | Adapter, quality and overrepresentation filtering |
| BWA | 0.7.19-r1273 | Reference indexing and alignment (BWA-MEM) |
| GATK | 4.3.0.0 | Read groups, duplicate marking, BQSR, variant calling and filtering |
| R (ggplot2, gplots, reshape, gsalib) | 4.5.3 | BQSR plots (AnalyzeCovariates) |
| Python (pandas, matplotlib) | 3.10 (2.3.3, 3.10.9) | Annotation statistics and plots |
| Ensembl VEP (web interface) | Release 2026-07 | Functional annotation |
| IGV-Web | igv.org/app | Visual inspection of alignments and variants |

**Table 2:** Dataset and reference used.

| Item | Details |
|---|---|
| Sample | NA12878 (BioSample SAMN00801888) |
| Study / run | PRJEB31736 (1000 Genomes high-coverage, New York Genome Center) / ERR3239334 |
| Sequencing | Illumina NovaSeq, 2 × 150 bp paired-end |
| Source file | NA12878.final.cram (15.8 GB), aligned to GRCh38 with decoy and HLA sequences |
| Reference genome | Broad Institute GRCh38, Homo_sapiens_assembly38.fasta (3.1 GB) |
| Region analysed | Chromosome 22 |

### Data source and read retrieval

The reference FASTA was indexed with samtools faidx and given a sequence dictionary with GATK CreateSequenceDictionary. Downloading the 15.8 GB CRAM file failed repeatedly over an unstable connection, so only the reads on chromosome 22 were fetched. A header request with curl confirmed that the EBI server accepts byte-range requests, after which samtools used the CRAM index to download only the blocks that cover chr22:

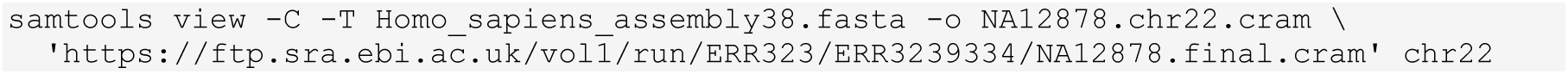

This produced a 168 MB CRAM file containing 10,104,487 records. The file was sorted by position so the two reads in each pair were very far apart and the first conversion to FASTQ only kept 21,940 pairs. The records were therefore grouped by read name with samtools collate before conversion:

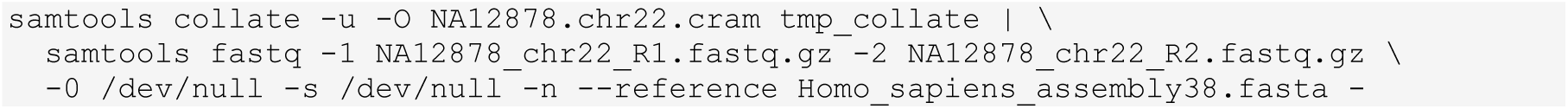

The output contained 1,541,984 read pairs in each file (114 MB and 119 MB). As explained in the Results (Read retrieval), this was later found to be a random subset of about 31% of the chr22 read pairs.

### Quality control and read cleaning

FastQC was run on both raw files and the reports were combined with MultiQC. A first round of trimming used Trimmomatic in paired-end mode with LEADING:3, TRAILING:3, SLIDINGWINDOW:4:15 and MINLEN:36. This command had no adapter-clipping step, and on review, adapters and overrepresented sequences were removed explicitly with fastp. fastp was therefore run on the original raw reads:

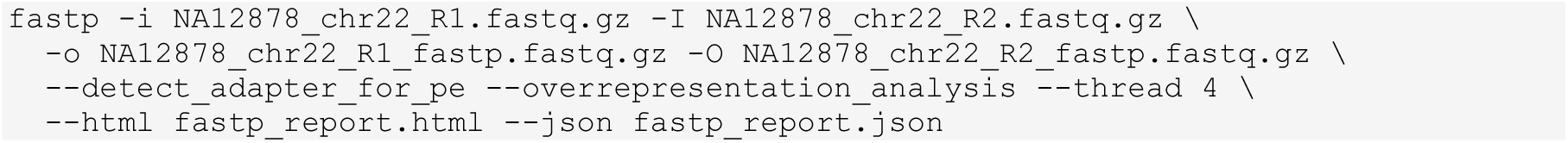

FastQC and MultiQC were then repeated on the fastp output, so that the raw, Trimmomatic and fastp results could be compared in one report. The fastp files were used in all later steps.

### Reference preparation and alignment

The total reference size was 3.1 GB. bwa index was run to create an index for the reference, which ran for approximately 49 minutes before being halted by the OS due to lack of memory. All reads were from chr22, so we extracted chr22 from the reference using samtools faidx. This created a FASTA file containing only chr22 (49 MB) with the original GRCh38 co-ordinates. This was indexed using samtools faidx. We also used GATK CreateSequenceDictionary and bwa index. The BWA index was created in 43 seconds. The clean reads were aligned to the reference (BWA-MEM) using 4 threads. Read groups were added using GATK AddOrReplaceReadGroups:

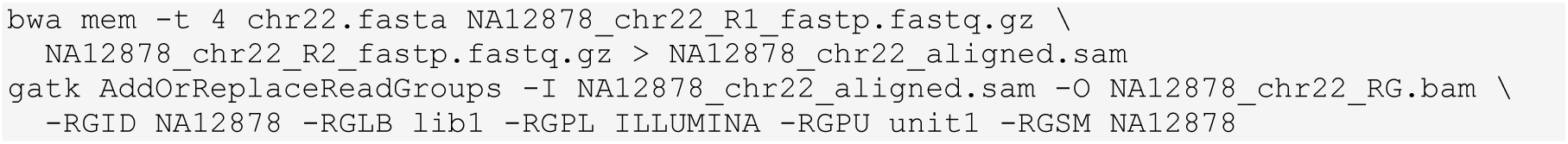

### BAM processing and validation

The BAM file was sorted and indexed with samtools (sort -@ 4 and index). PCR and optical duplicates were marked with GATK MarkDuplicates (they were flagged rather than removed) and the output was indexed again. The number of reads with the duplicate flag was determined via samtools view -c -f 1024, coverage statistics for the chromosome were generated with samtools coverage and the alignments were checked in IGV-Web against the hg38 genome.

### Base quality score recalibration

BQSR needs catalogues of known variants. The standard dbSNP 138 file for hg38 is an uncompressed 10.95 GB VCF that cannot be queried by region over the network, so it was not used. Instead, three compressed and indexed resources from the Broad Institute public bucket (gcp-public-data--broad-references/hg38/v0) were streamed for chr22 only with bcftools view -r chr22 (Table 3). The 1000 Genomes phase 1 high-confidence SNP set took the place of dbSNP as the SNP resource.

**Table 3:** Known-variant resources used for BQSR (chr22 extracts)

| **Resource** | **Type** | **chr22 records** | **Size** |
|---|---|---|---|
| 1000G_phase1.snps.high_confidence.hg38 | SNPs | 328,813 | 20 MB |
| Mills_and_1000G_gold_standard.indels.hg38 | Indels | 18,679 | 300 KB |
| Homo_sapiens_assembly38.known_indels | Indels | 26,708 | 805 KB |

BaseRecalibrator was run on the duplicate-marked BAM with the three resources and a 4 GB Java heap, and ApplyBQSR wrote the recalibrated BAM. A second BaseRecalibrator pass on the recalibrated BAM produced an ‘after’ table, and AnalyzeCovariates compared the two tables. Because AnalyzeCovariates draws its plots with R, R 4.5.3 with the packages ggplot2, gplots, reshape and gsalib was installed in the separate r-plots environment and added to the PATH for that single command.

### Variant calling and hard filtering

Variants were called using GATK HaplotypeCaller on the recalibrated BAM (native pair HMM threads set to 4, 4 GB heap), resulting in a compressed raw VCF. SNPs were extracted using SelectVariants (--select-type-to-include SNP). Indels were selected together with mixed sites (where a SNP and an indel occur at the same position) using --select-type-to-include INDEL and MIXED. Without the MIXED option, 68 of these records were missing from both the SNP and indel sets. Each of the two sets was then filtered using VariantFiltration with the GATK-recommended starting thresholds as listed in Table 4. The two filtered VCFs were then combined using MergeVcfs. Variants that failed filtering were simply annotated with a failure in the FILTER column rather than being removed. Variant counts and Ti/Tv ratios were then derived from the resulting VCF using bcftools stats.

**Table 4:** Hard-filter thresholds applied with GATK VariantFiltration.

| **Annotation** | **Meaning** | **SNP fails if** | **Indel fails if** |
|---|---|---|---|
| QD | Variant confidence normalised by depth | < 2.0 | < 2.0 |
| QUAL | Phred-scaled call confidence | < 30.0 | < 30.0 |
| FS | Strand bias (Fisher’s exact test) | > 60.0 | > 200.0 |
| SOR | Strand bias (symmetric odds ratio) | > 3.0 | > 10.0 |
| MQ | Root-mean-square mapping quality | < 40.0 | not applied |
| MQRankSum | Mapping quality, alt vs ref reads | < −12.5 | not applied |
| ReadPosRankSum | Position of the alt allele in reads | < −8.0 | < −20.0 |

### Functional annotation

Only variants with FILTER = PASS were annotated (bcftools view -f PASS). Ensembl names chromosomes without the ‘chr’ prefix, so chr22 was renamed to 22 with bcftools annotate --rename-chrs. The file was submitted to the Ensembl VEP web interface (release 2026-07) with these settings: human GRCh38.p14, GENCODE 50 transcript set, HGVS notation, AlphaMissense predictions, gnomAD genomes v4.1.1 allele frequencies, and phenotype and disease associations (ClinVar fields were present in the output). The results were downloaded in VCF and tab-separated (TSV) formats. For viewing in IGV, the annotated VCF was re-compressed with bgzip and indexed with bcftools.

### Summary statistics and visual inspection

Each variant allele generates one row in VEP per overlapping transcript, and hence, the original 780,634 rows in the TSV file were condensed by running a compact Python code snippet (pandas 2.3.3, matplotlib 3.10.9). Rows are summarised based on their genomic location, reference, and alternative alleles while keeping the most severe effect per allele. This ranking of severity is done on the basis of IMPACT (HIGH > MODERATE > LOW > MODIFIER), where the canonical transcript is preferred in case of equal impact in two rows. A variant is classed as a known variant if it has an rsID. In the current VEP output, there are no values in the Existing_variation column; hence, rsIDs have been obtained from the gnomAD genomes annotation. The frequency of the alleles has been described as common (≥ 5%), low frequency (1-5%), rare (< 1%) or not in gnomAD. Significance from ClinVar has been derived on the basis of a pattern that does not include ‘conflicting classifications of pathogenicity’.

The code further identified 63 variants as having either HIGH impact or being predicted by AlphaMissense to be pathogenic. Four such variants have been chosen to represent different categories and levels of confidence and were analysed in IGV-Web along with the recalibrated BAM and VCF.

## Results

### Read retrieval

Streaming just chromosome 22 brought the download down from 15.8 GB to 168 MB. The chr22 CRAM had 10,104,487 records and the final FASTQ files had 1,541,984 pairs (3.08 million reads). When the working directory was subsequently cleaned up there were still the 0020 to 0063 temp files from samtools collate present. Collate works by taking the reads and distributing them into 64 temp files by read name before processing them sequentially one at a time. The fact that only 20 of the 64 temp files had been processed (20/64 = 31%) is very close to the proportion of reads that were recovered (3.08 million of about 10.1 million records).

As collate assigns the pairs to its temporary files by read name rather than by position, the pairs that are retained form an effectively random sample spread right across the chromosome. This is what gives the even coverage that we see in IGV. All the subsequent results are internally consistent for this subset; the effective depth of 8.6x (see Alignment and BAM processing) is a result of this step and not of the original data which had been sequenced to around 30x. The issue was not spotted until after the analysis was complete and is listed as one of the limitations in the Limitations section.

### Quality control

The raw reads were of high quality (Figure 2). Mean Phred scores stayed between about 28 and 30 along the whole 150 bp read with no drop at the 3′ end, duplication was 4.2–4.3%, GC content was 47%, and FastQC found adapter sequence in no more than about 0.13% of reads at any position. The only warning concerned per-sequence GC content, which is expected: FastQC compares the reads with a whole-genome model, and chr22 is richer in GC than the genome average.

**Figure 2:**
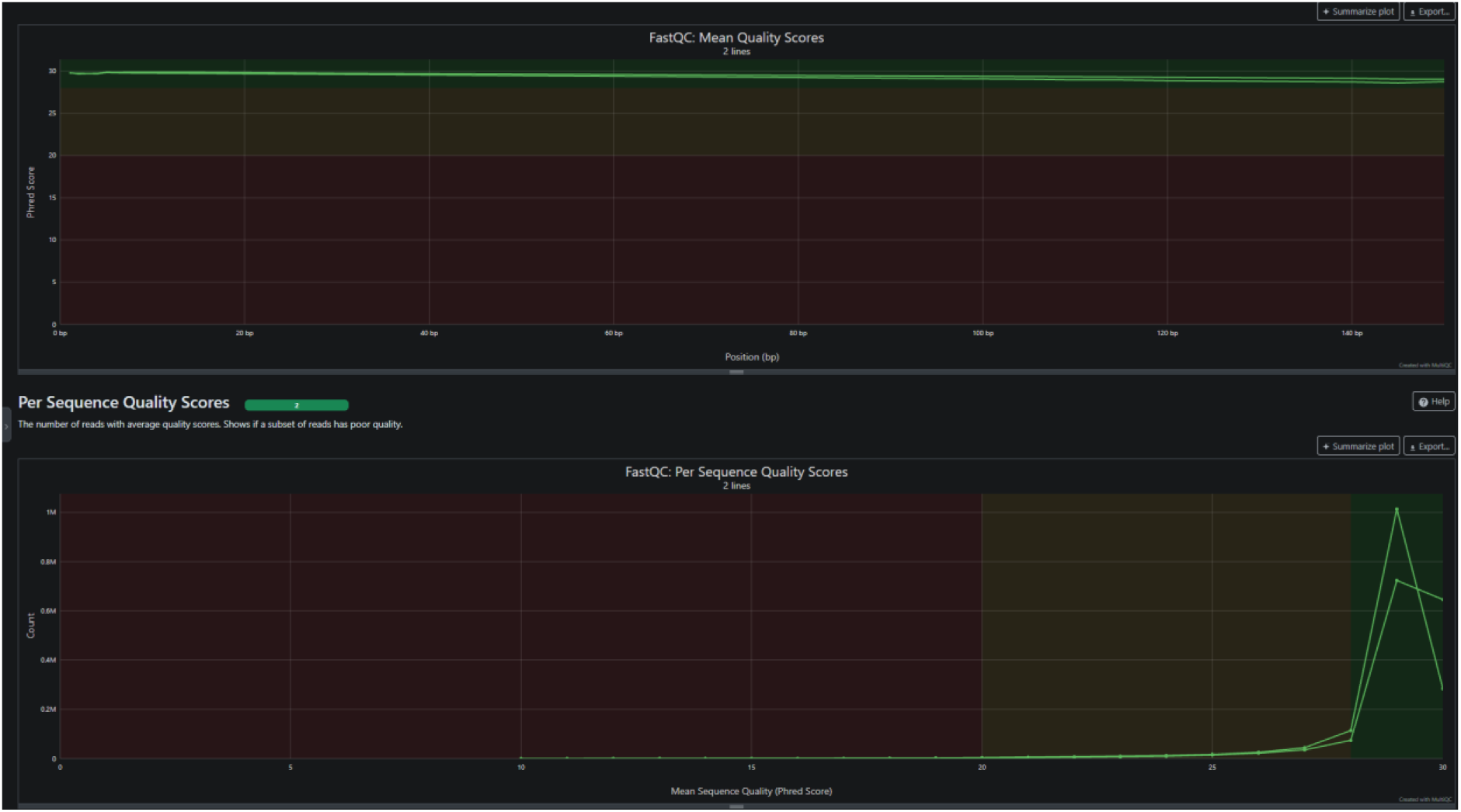
MultiQC summary of the raw reads: mean quality along the read (top) and per-sequence quality scores (bottom).

Trimmomatic kept both reads for 1,528,266 of 1,541,984 pairs (99.11%); 10,703 forward-only reads (0.69%), 2,778 reverse-only reads (0.18%) and 237 pairs (0.02%) were dropped. The fastp results are summarised in Table 5. fastp detected a small amount of TruSeq adapter (about 0.27% of reads) and removed it. The overrepresented sequences it reported were simple repeats such as poly-A and AC dinucleotide runs, each in fewer than 0.15% of reads, and it estimated the insert size peak at about 268 bp.

**Table 5:**
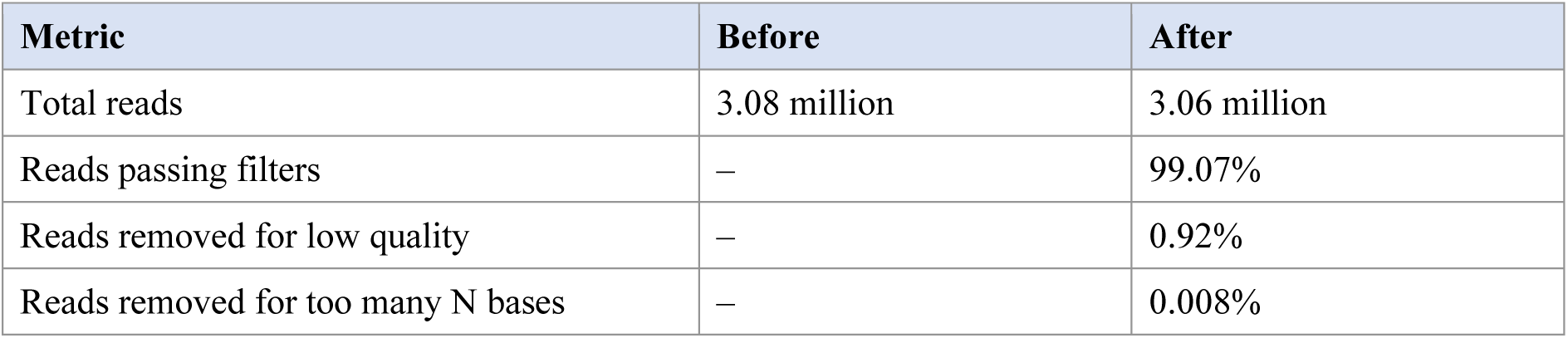
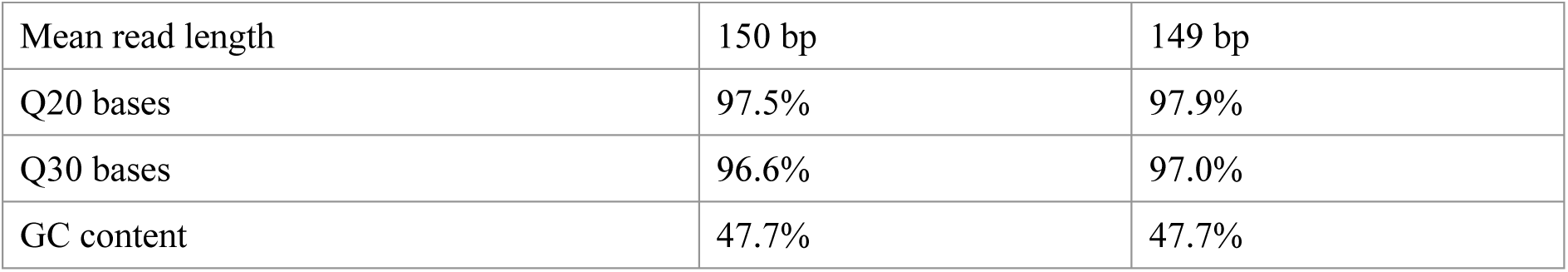
Read statistics before and after fastp.

The trimmed reads passed every FastQC module except two expected warnings (Figure 3): GC content, for the reason given above, and sequence length distribution, because trimmed reads no longer all have the same length. The same pattern was seen for the fastp output in the combined six-sample MultiQC report. Since the raw data were already clean, the cleaning steps changed the read set only slightly, but they made sure that adapter removal was carried out and documented.

**Figure 3:**
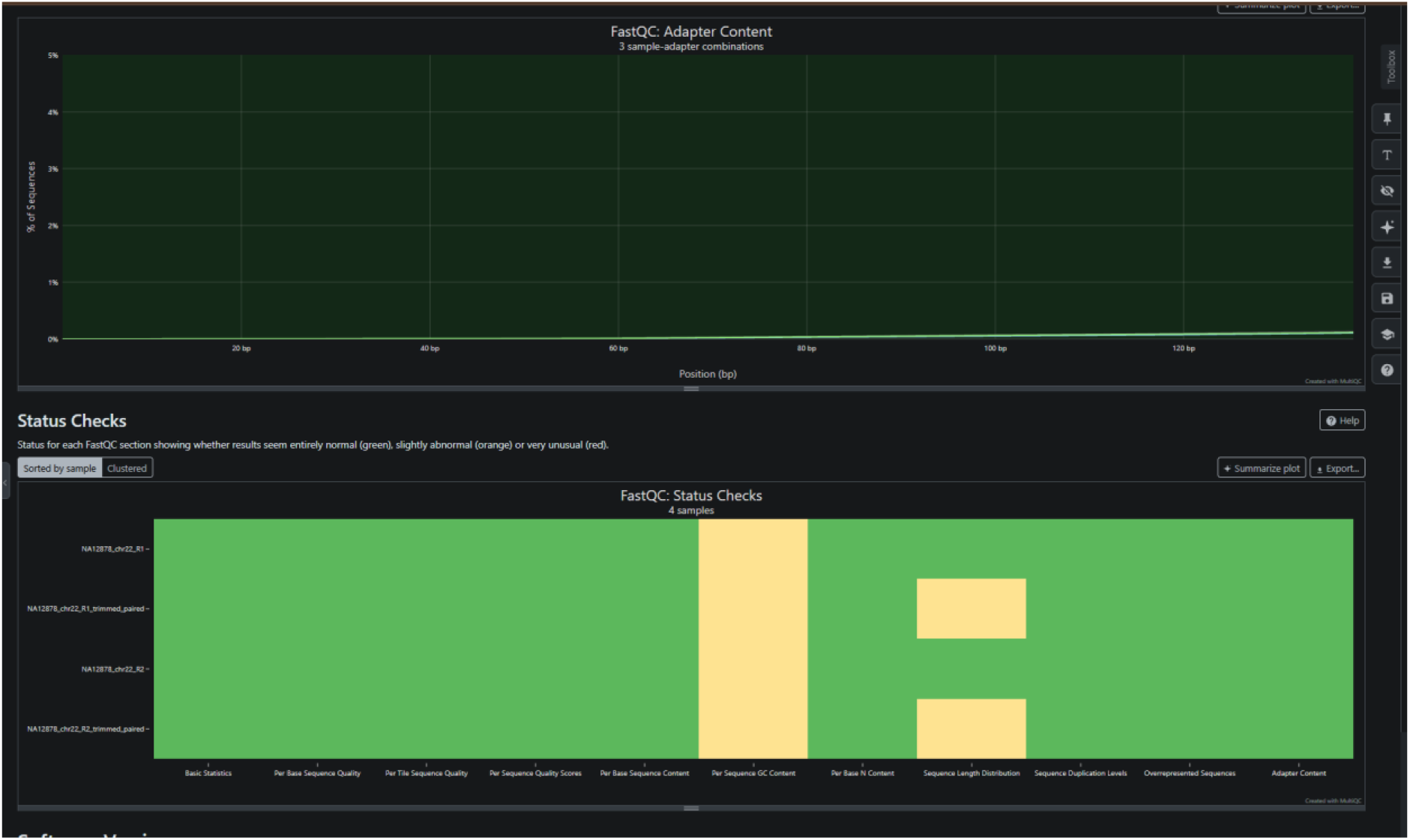
Adapter content and FastQC status checks for the raw and Trimmomatic-trimmed reads (MultiQC). Green = pass, orange = warning.

### Alignment and BAM processing

BWA-MEM aligned the reads to chr22 in approximately 2.6 minutes to produce a 1.3 GB SAM file which became a 385 MB BAM file after read groups were added and 149 MB after sorting. MarkDuplicates identified 104,691 of the 3,058,443 records (3.4%) as duplicates, including 7,512 optical duplicate pairs, and samtools confirmed the same number of flagged reads. The coverage statistics are in Table 6.

**Table 6:**
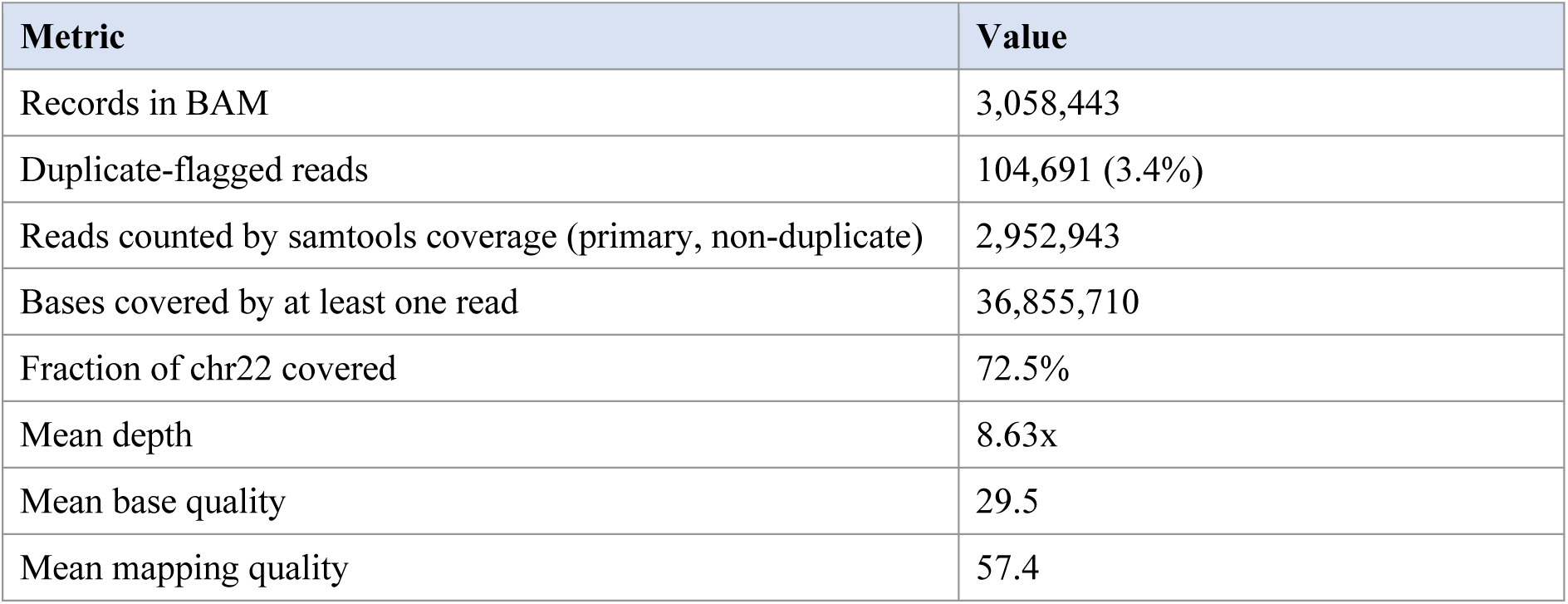
Alignment and coverage statistics for the duplicate-marked BAM.

About 27.5% of chr22 had no reads, which corresponds mostly to the unsequenced short arm and the centromeric gap at the start of the chromosome. A mean mapping quality of 57.4, close to the maximum of 60, shows that most reads were placed unambiguously. In IGV (Figure 4) the reads were stacked evenly on both strands, scattered single mismatches were rare, and columns of consistent mismatches marked likely true variants.

**Figure 4:**
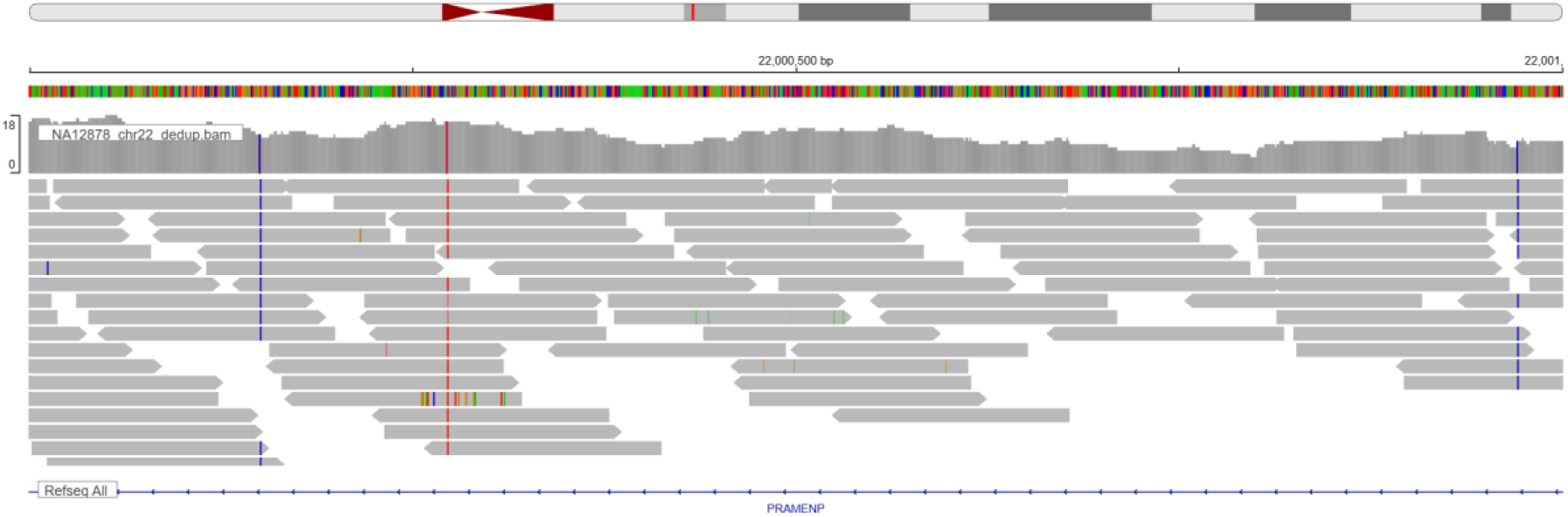
IGV view of the duplicate-marked BAM at chr22:22,000,000–22,001,000 (PRAMENP region).

### Base quality score recalibration

BaseRecalibrator used 2,881,860 reads after excluding duplicate-flagged and zero-MAPQ reads, and the recalibrated BAM (302 MB) contained exactly the same 3,058,443 records as its input. The data were already well calibrated: the overall reported quality was Q25.1 against an empirical Q25.0, and 97% of the 424 million observed bases had a reported quality of Q30 (empirical Q29). Smaller biases were present. Bases reported as Q20 behaved like Q17, quality in the middle of reads was about one point too optimistic with a dip of about 3.5 points near the start of reads, and a few sequence contexts (AC, CT and GC) were off by 1–2.5 points. After recalibration these deviations moved close to zero (Figure 5), with a slight positive overshoot in the middle of reads.

**Figure 5:**
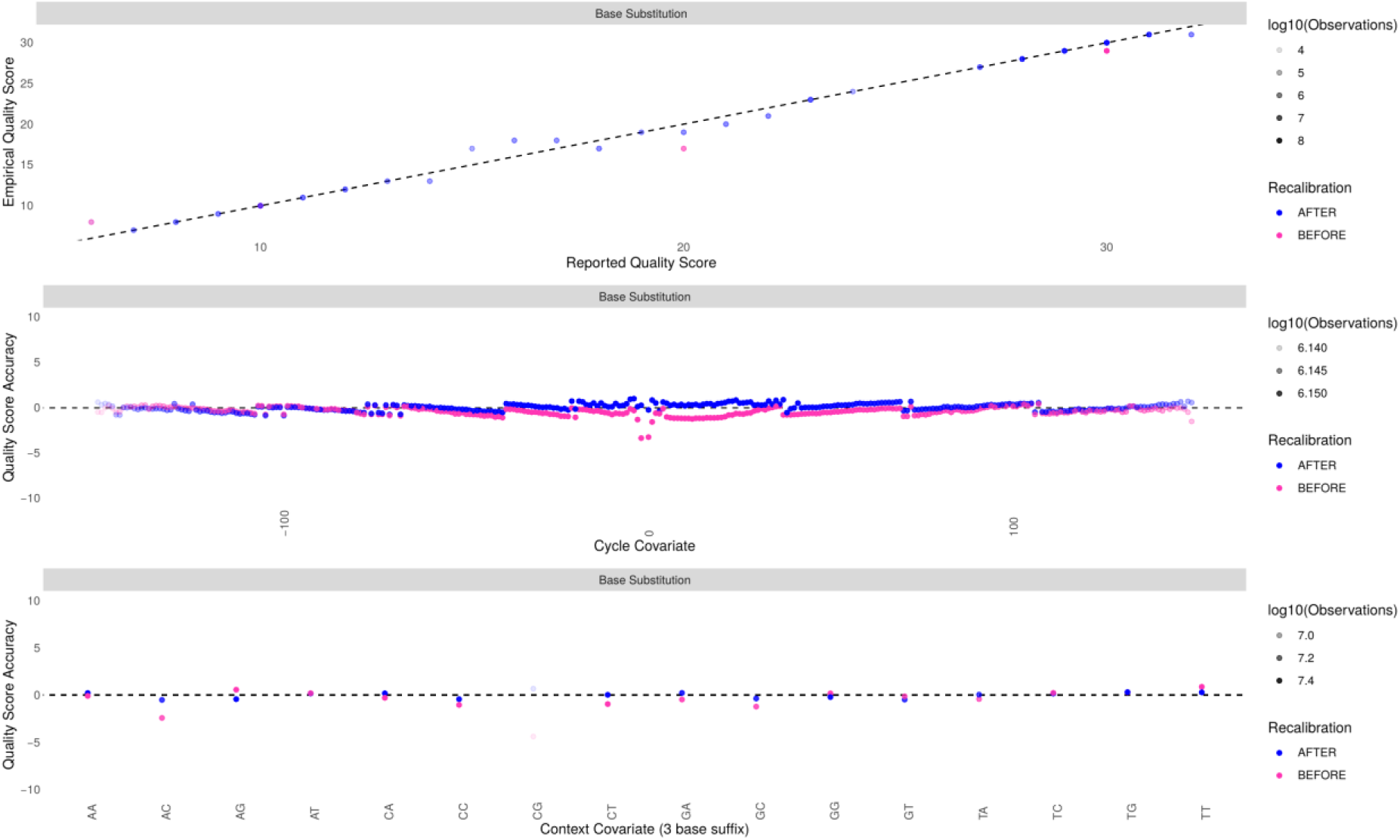
Base quality before (pink) and after (blue) recalibration: reported versus empirical quality (top), quality accuracy by cycle (middle) and by sequence context (bottom), from GATK AnalyzeCovariates.

The magnitude of the corrections is very much in keeping with the recent Illumina data (the quality scores are binned and generally good). Using the 1000 Genomes high-confidence SNPs rather than dbSNP meant that far fewer known sites were masked, so some real variants may have been counted as errors. Given that the corrections were so small the impact on the final calls is probably negligible.

### Variant calling and hard filtering

HaplotypeCaller ran for 38.3 minutes and generated 81,599 variant records. bcftools reported 69,591 SNPs and 12,056 indels (with 637 multiallelic sites), so a few records are in both sets. The SNP set contained 69,543 records and the indel+mixed set 12,056, which gives the full 81,599. After hard filtering, 73,570 variants (90.2%) passed (Table 7).

**Table 7:** Outcome of hard filtering.

| **FILTER value** | **Variants** |
|---|---|
| PASS | 73,570 (90.2%) |
| MQ40 | 3,483 |
| SOR3 | 2,648 |
| QD2 | 1,236 |
| MQ40;SOR3 | 465 |
| MQ40;QD2 | 105 |
| QD2;SOR3 | 88 |
| MQ40;QD2;SOR3 | 4 |
| Total | 81,599 |

The Ti/Tv ratio for the PASS SNPs was 2.04 (41,423 transitions and 20,294 transversions), which is in the expected range for whole-genome data and a good indicator that the PASS set is not skewed towards false positives. No variant failed the QUAL filter, as HaplotypeCaller by default does not output calls below a confidence of 30. The most common reason for failure was low mapping quality (MQ40), which is indicative of reads in repetitive/duplicated sequence, with strand bias (SOR3) being the next most common.

### Functional annotation

VEP annotated all 73,570 PASS records, generating 780,634 transcript-level rows which the Python script then collapsed down to 74,061 variant alleles. The difference from the 73,570 is due to the multi-allelic sites where each alternative allele is individually counted. Table 8 contains the summary of the results.

**Table 8:** Summary of VEP annotation (per variant allele, most severe consequence)

| **Category** | **Count** |
|---|---|
| Variant alleles annotated | 74,061 |
| SNVs / indels and other | 61,707 / 12,354 |
| Known (rsID) / novel | 73,235 (98.9%) / 826 (1.1%) |
| Coding in at least one transcript | 734 alleles in 270 genes |
| Missense (most severe consequence) | 407 |
| Impact HIGH / MODERATE / LOW / MODIFIER | 57 / 428 / 479 / 73,097 |
| AlphaMissense (missense) | 6 likely pathogenic, 5 ambiguous, 244 likely benign, 152 not scored |
| With a ClinVar record | 1,029 (968 benign) |
| ClinVar pathogenic or likely pathogenic | 0 |

As expected for a whole genome, most variants lay outside coding sequence: 34,947 were intergenic and 26,597 intronic (Figures 6 and 7). Almost 99% of alleles already had an rsID and 89% were common in gnomAD (Figure 8); only 857 alleles (1.2%) were absent from gnomAD. This is the pattern expected for a healthy and extensively studied individual, and it supports the reliability of the PASS calls. ClinVar held records for 1,029 of the variants, almost all classified as benign or likely benign, with 15 of uncertain significance and nine labelled as drug response. None was pathogenic or likely pathogenic, which again fits a healthy reference individual.

**Figure 6:**
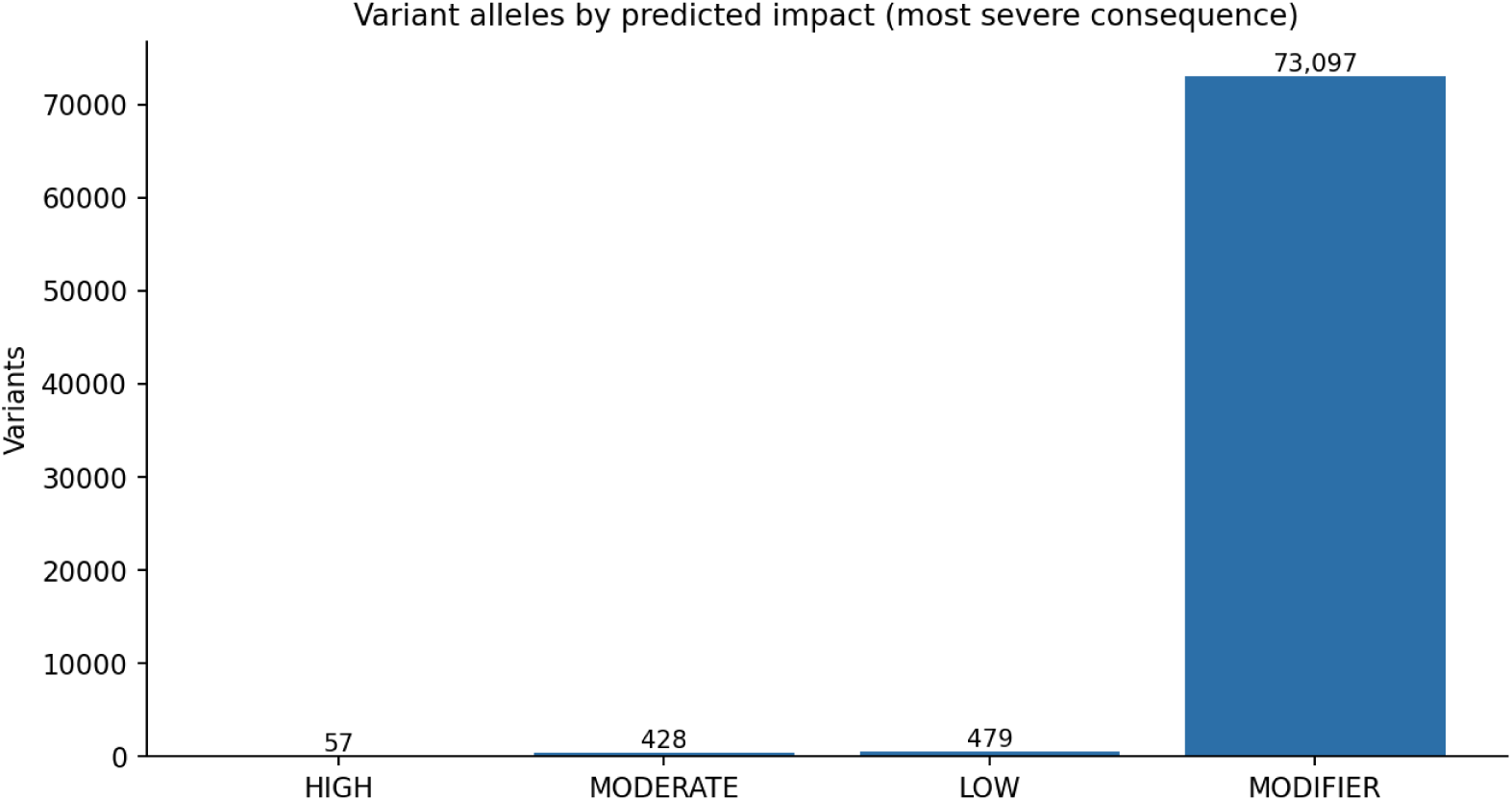
Number of variant alleles in each VEP impact class (most severe consequence per allele).

**Figure 7:**
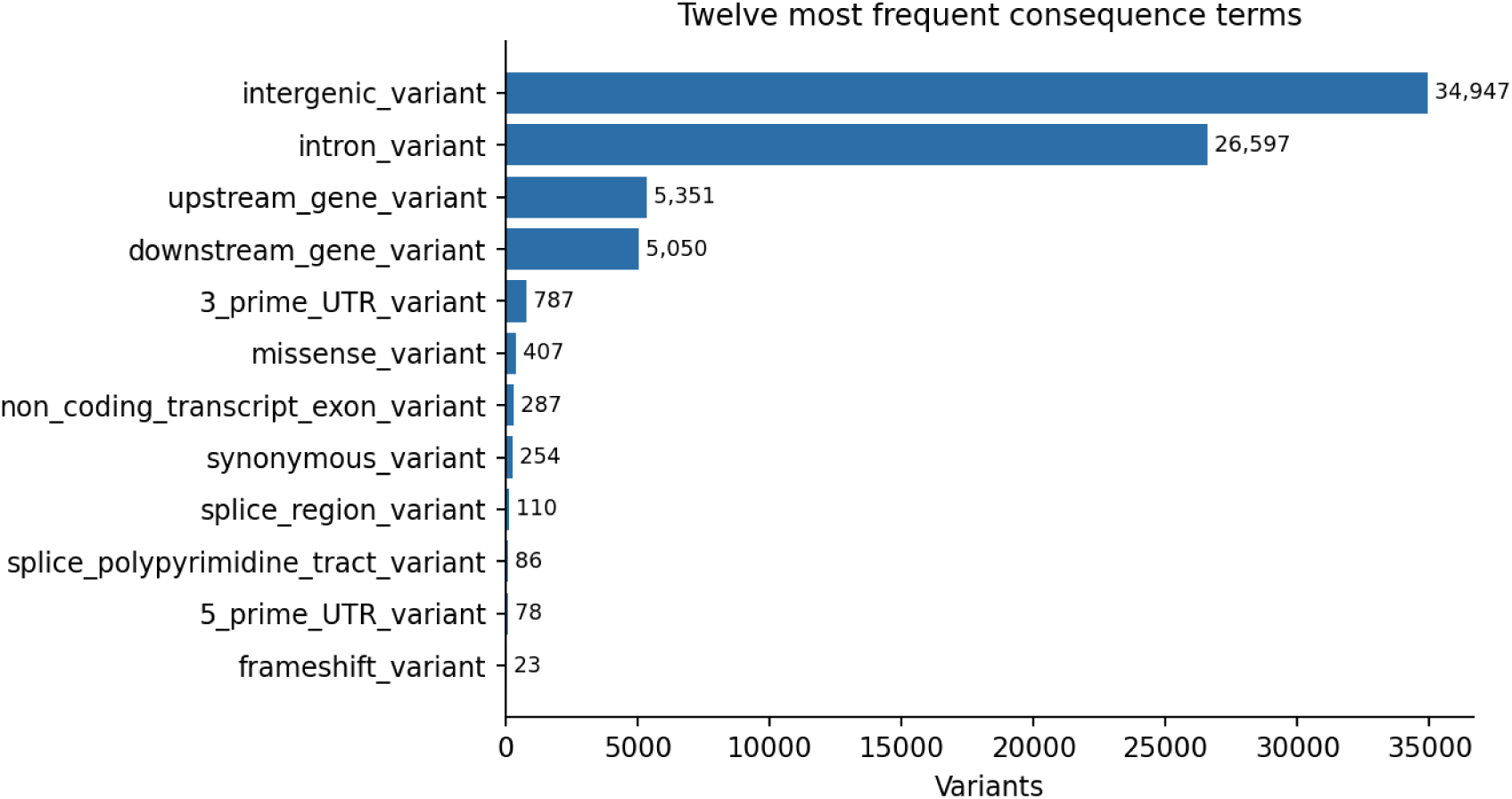
The twelve most frequent consequence terms among the annotated variant alleles.

**Figure 8:**
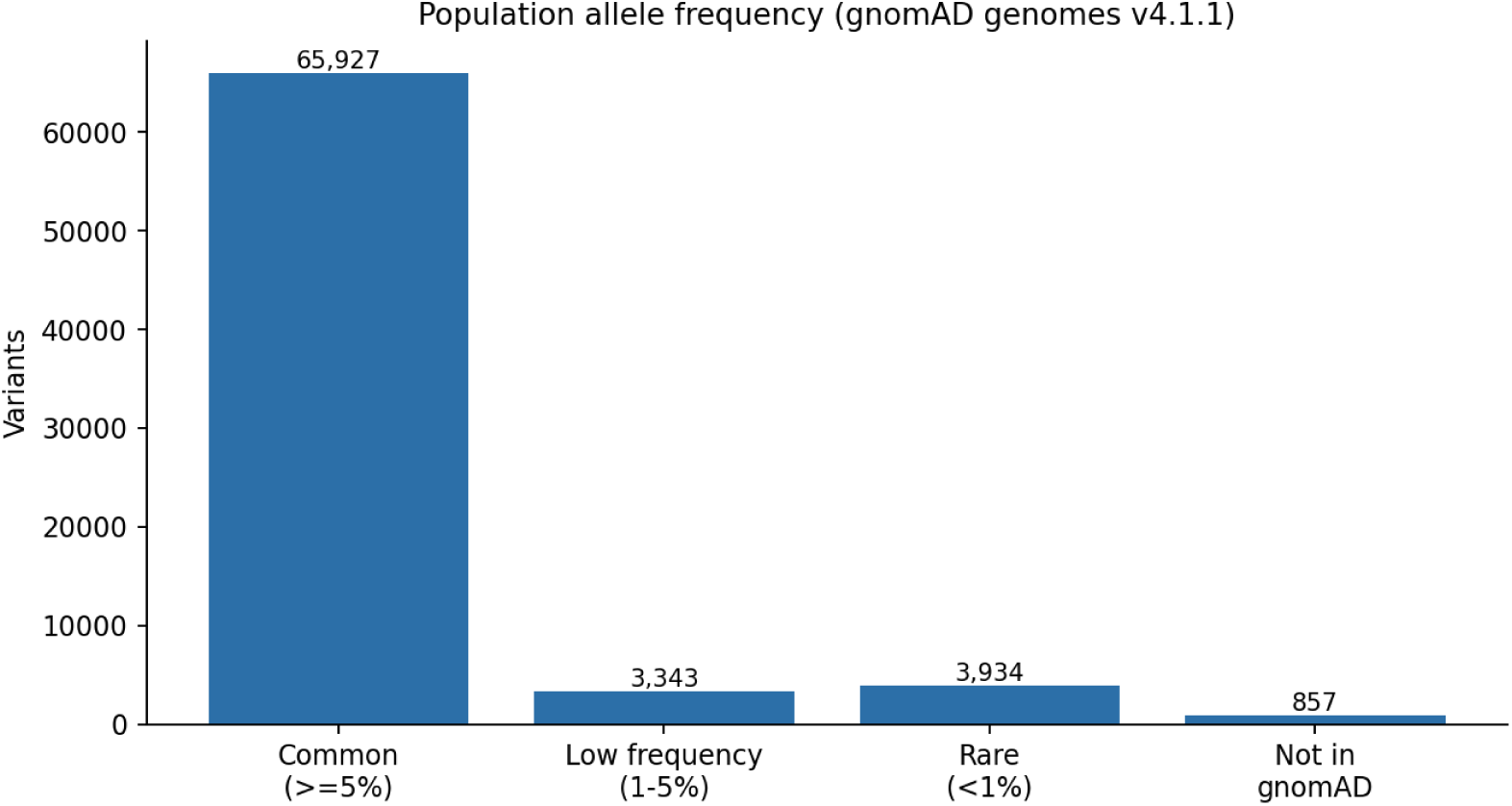
Population allele frequency of the annotated variants in gnomAD genomes v4.1.1.

Two points call for caution. First, missense variants outnumbered synonymous ones (407 against 254), and the genes with the most coding variants included PKDREJ, GSTT4, the immunoglobulin lambda genes IGLV5-48 and IGLV4-69, CYP2D7, CYP2D6 and POTEH, several of which belong to families of very similar genes. Some of these calls may come from reads that belong to paralogous copies, a risk that the chr22-only reference increases. Second, the script reports the most severe consequence across all transcripts, so a variant that is intronic in the main transcript but coding in an alternative one is counted as coding. The HPS4 variant in the section on manual inspection of selected variants is one such case, so the count of 57 HIGH-impact alleles should be read as an upper limit.

### Manual inspection of selected variants

Four variants from the candidate list were examined in IGV (Table 9).

**Table 9:** Variants inspected in IGV.

| **Gene** | **Position (GRCh38)** | **Change** | **Annotation** | **Read evidence** | **gnomAD AF** | **Assessment** |
|---|---|---|---|---|---|---|
| RTCB | chr22:32,408,189 | G>T (rs73168800); NM_014306.5:c.226C>A | Missense; AlphaMissense likely pathogenic | 10/17 alt (6+/4−); heterozygous | 0.000013 | Well supported |
| CYP2D6 | chr22:42,128,945 | C>T (rs3892097); NM_000106.6:c.506-1G>A | Splice acceptor; defines CYP2D6*4 | 4/14 alt (2+/2−) | 0.14 | Supported; low allele fraction |
| PRODH | chr22:18,922,938 | 1-bp deletion (CA>C) | Frameshift in one transcript | 5/5 reads; homozygous call | Absent | Low confidence |
| HPS4 | chr22:26,466,075 | G>A (rs3747129); NM_022081.6:c.706+151C>T | Stop-gained in an alternative transcript; intronic in NM_022081.6 | 4/7 alt (2+/2−); heterozygous | 0.12 | Real; limited impact |

The RTCB variant had the strongest support (Figures 9 and 10). Ten of 17 reads carried the T allele, on both strands; the reads had high mapping quality and the variant lay in the middle of reads rather than at their ends. Its VCF record showed QUAL 283.64, QD 16.68, FS 1.957, SOR 0.956 and MQ 60, all well inside the filter limits, and an allele frequency of 0.5, consistent with a heterozygous call. The variant is very rare in gnomAD and AlphaMissense predicts it to be likely pathogenic, but a single heterozygous variant predicted to be damaging, in a healthy person, is a prediction and not a clinical finding.

**Figure 9:**
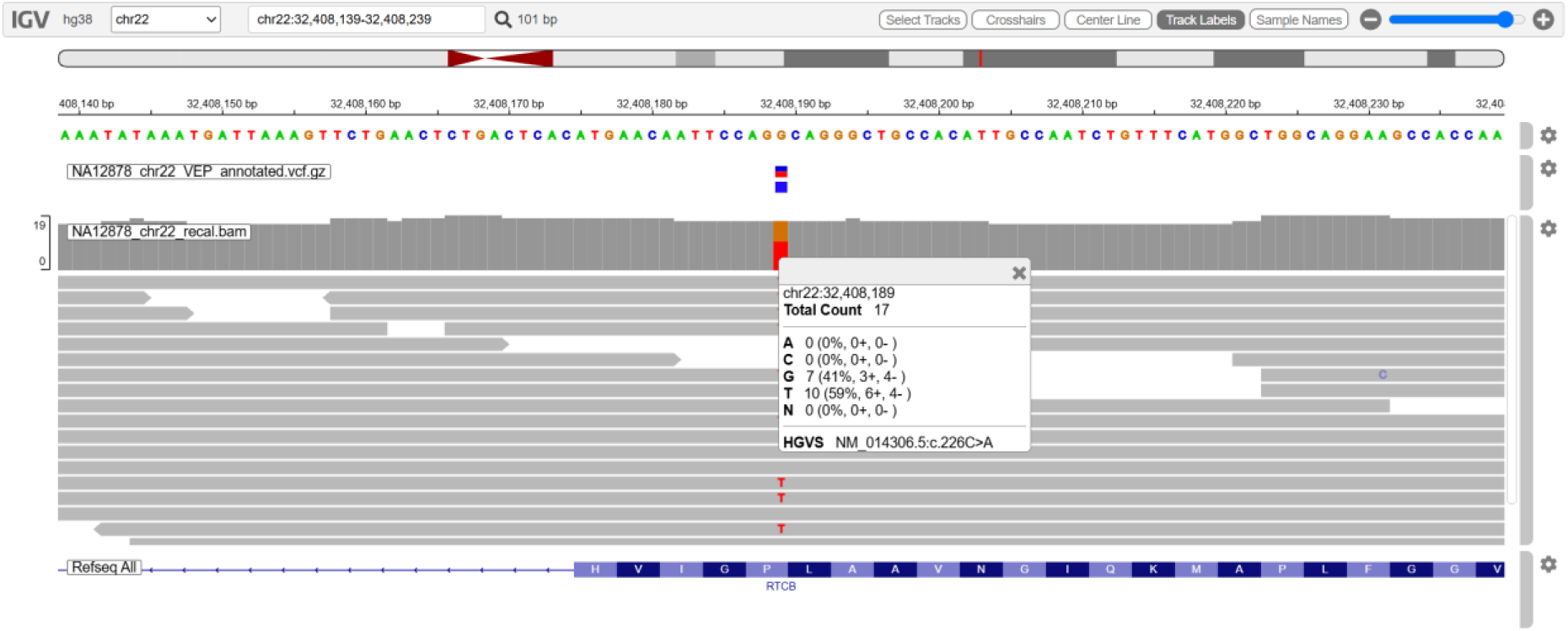
RTCB missense variant (chr22:32,408,189 G>T) in IGV with the annotated VCF, the recalibrated BAM and the base counts at the variant position.

**Figure 10:**
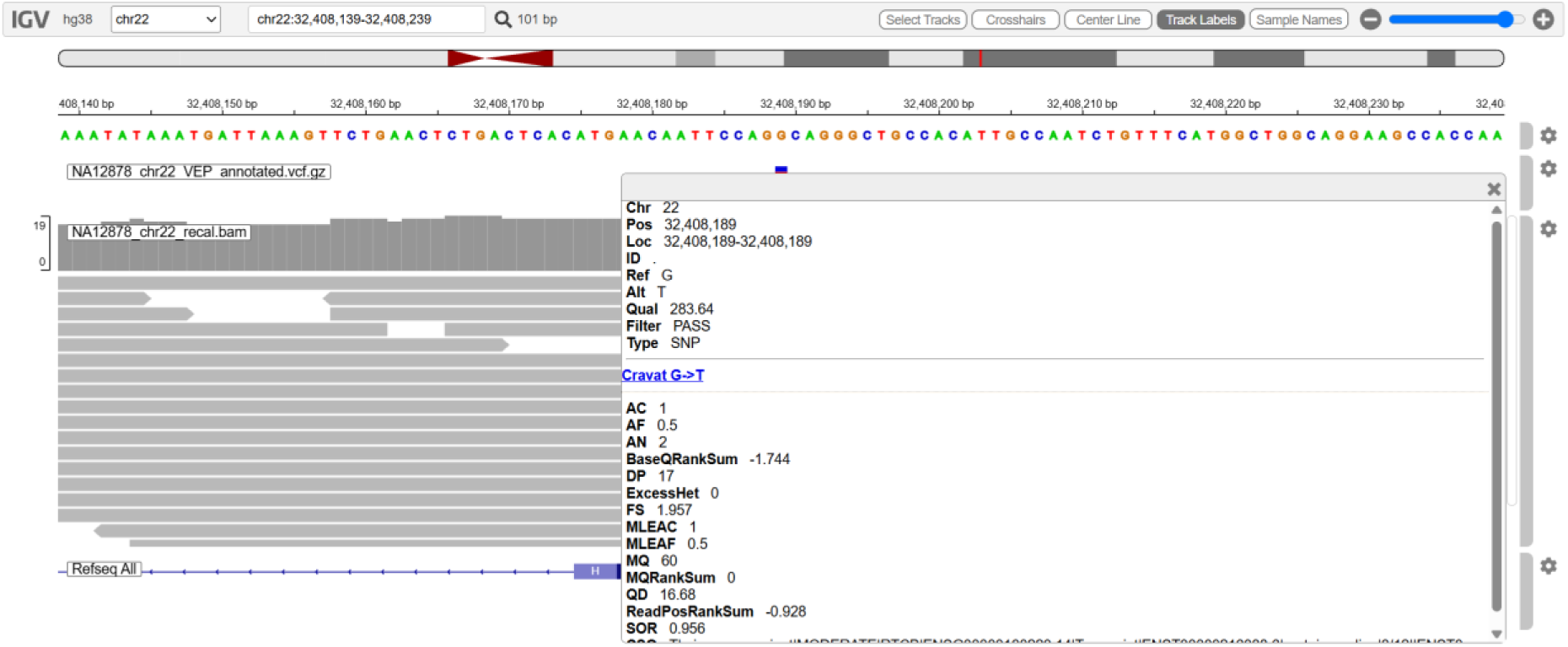
VCF record of the RTCB variant in IGV, showing the quality annotations and the VEP annotation (CSQ) field.

At the CYP2D6 splice acceptor site (Figure 11), 4 of 14 reads had the variant, again on both strands. IGV linked the position to the ClinVar entry NM_000106.6:c.506-1G>A (the splice defect that defines the CYP2D6*4 allele) and the frameshift that defines CYP2D6*3 (rs35742686) was also one of the HIGH-impact calls. GeT-RM lists NA12878 as having the consensus genotype *3/(*68)+*4 [25], so the presence of both defining variants is in accord with the published result. The allele fraction of 29% is less than the 50% we would expect at a heterozygous site. With only 14 reads, this could easily be due to chance and the presence of the extra CYP2D6–CYP2D7 hybrid copy (*68) and the high degree of sequence similarity between the two genes may also have an effect on the read counts. The hybrid itself cannot be resolved by short-read SNV calling, which is why tools such as Cyrius were developed [24].

**Figure 11:**
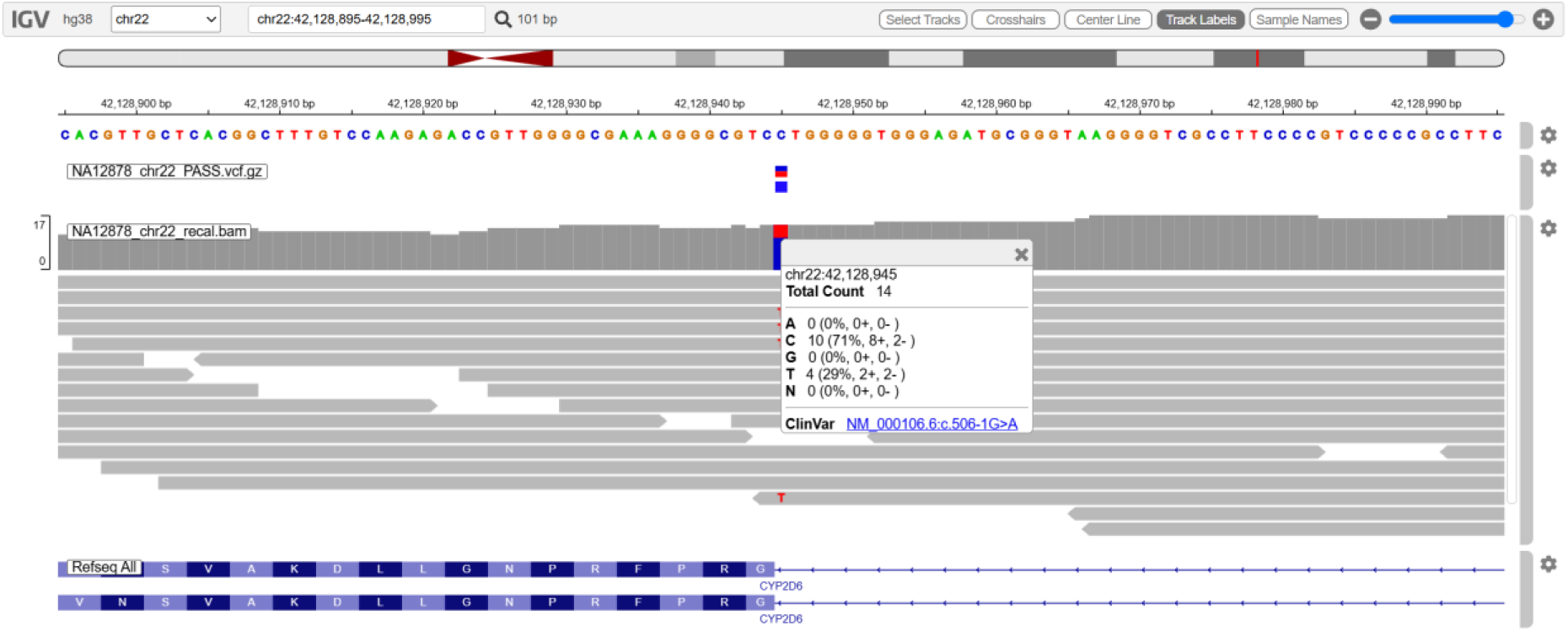
CYP2D6 splice-acceptor variant c.506-1G>A (CYP2D6*4) at chr22:42,128,945 in IGV.

The PRODH deletion (Figure 12) was present in all five reads covering the position and was called homozygous, but the depth was low, the variant is absent from gnomAD and it lies in the duplication-rich 22q11 region. In the RefSeq transcripts shown in IGV it falls just outside the coding exon, so the frameshift label probably comes from a different GENCODE transcript. It is best treated as a low-confidence call that would need confirmation by another method.

**Figure 12:**
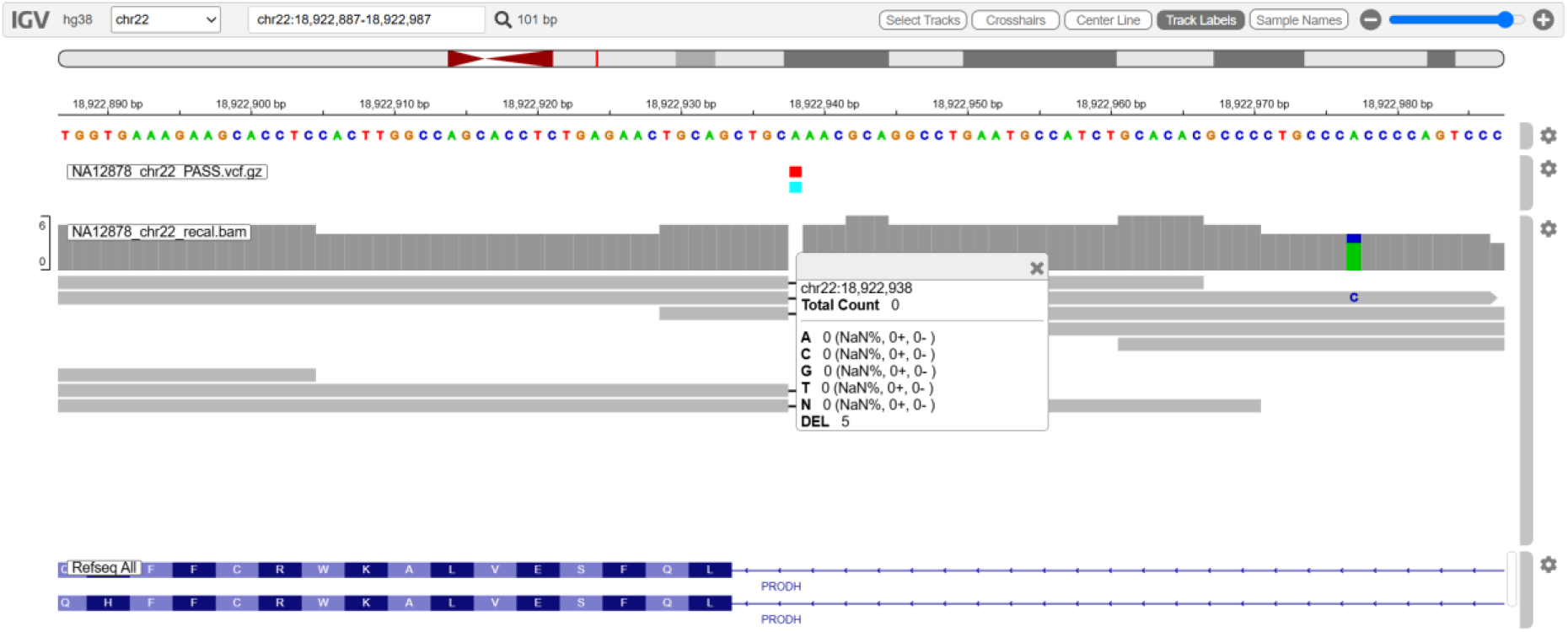
One-base deletion in PRODH at chr22:18,922,938 in IGV (DEL = 5 reads, low depth).

The HPS4 variant (Figure 13) was clearly heterozygous, with 4 of 7 reads on both strands. IGV named it NM_022081.6:c.706+151C>T, an intronic position in that transcript, while in an alternative transcript the same change creates a stop codon. This explains why VEP flagged it as HIGH impact even though it is common (gnomAD frequency 0.12) and probably of little functional consequence.

**Figure 13:**
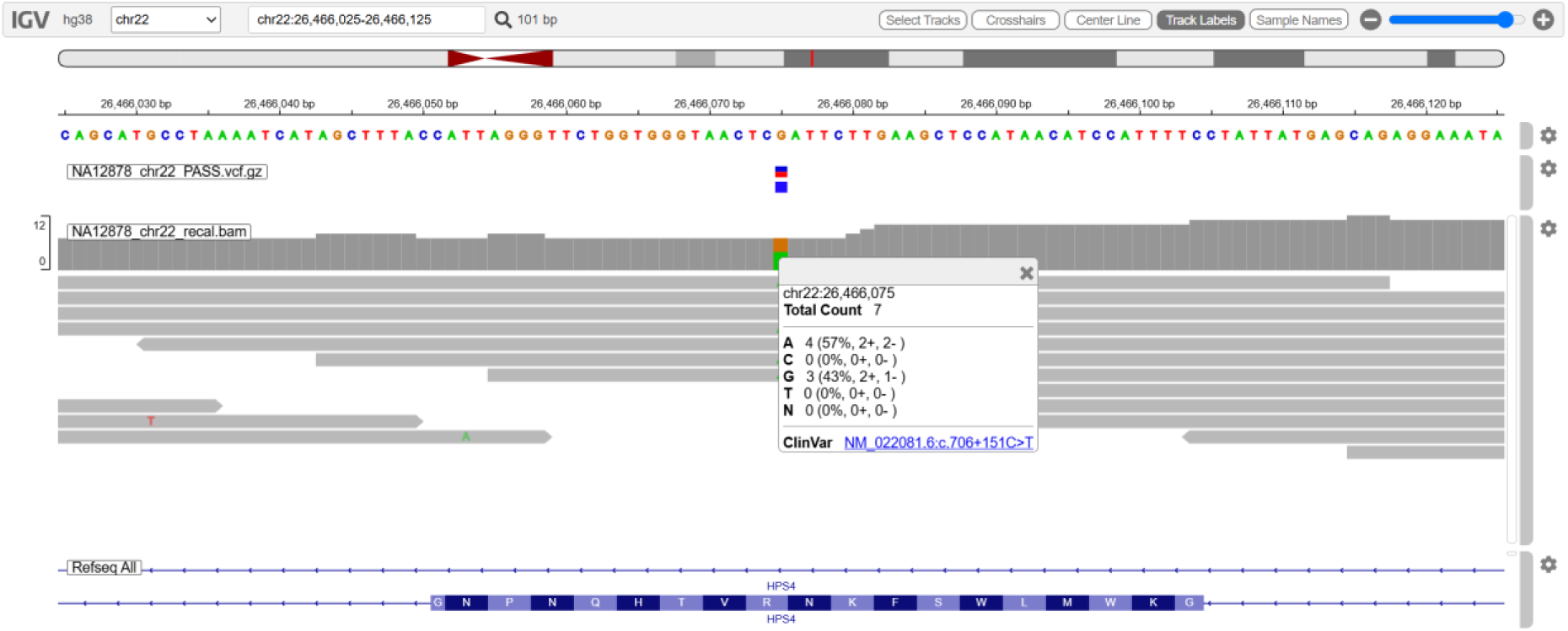
HPS4 variant at chr22:26,466,075 in IGV: intronic in NM_022081.6 (upper gene track) but within a coding exon of an alternative transcript (lower track).

Taken together, these checks showed that calls with good depth, balanced strand support and high mapping quality looked convincing in the reads, whereas calls in low-coverage or duplicated regions deserved caution even when they had passed the filters.

## Discussion

A full germline variant-calling workflow was developed and executed on chromosome 22 of NA12878 using a laptop that had 8 GB of memory. Each step in the GATK Best Practices was carried out and verified. The reads for chr22 were obtained without having to download the entire genome, the quality of the reads was checked and the adapter sequences were removed using fastp, the reads were then aligned and transformed into an analysis-ready BAM file, the base qualities were recalibrated, and the 81,599 raw calls were reduced by hard filtering to 73,570 PASS variants with a Ti/Tv ratio of 2.04. When the variants were annotated with VEP, a variant profile typical of a healthy and well-studied genome was observed, and examination in IGV confirmed the read support for the variants that had been selected, including the CYP2D6*3 and *4 defining variants which correspond to the published genotype of NA12878. On the way, a number of practical problems were dealt with, such as those concerning memory limitations when indexing, instability of the network connection, errors in pairing during FASTQ conversion, and a type of mixed variants which were lost during the filtering process. Moreover, when the work was reviewed it became clear that only about 31% of the read pairs had been carried forward. Identifying and recording these issues was just as important a part of the project as actually running the tools.

### Limitations

- **Effective depth.** The incomplete collate step kept a random ∼31% of the chr22 read pairs, giving a mean depth of 8.6x instead of about 30x. Sensitivity, particularly for heterozygous variants, is therefore lower than the source data would allow.
- **Chromosome-only reference.** Reads that truly come from similar sequence on other chromosomes had nowhere else to align. This can inflate mapping quality and allow paralog-derived false positives to pass the MQ filter.
- **Known sites for BQSR.** The 1000 Genomes high-confidence SNPs replaced dbSNP 138, which was too large to download in this setting.
- **Filtering.** The hard-filter thresholds are GATK’s generic starting values and were not tuned for this dataset, and mixed SNP–indel sites were filtered with the indel thresholds.
- **Consequence counting.** Reporting the most severe consequence across all transcripts overstates the number of HIGH-impact variants; counting with MANE Select transcripts only would be more conservative.
- **No formal benchmark.** The calls were not compared with the Genome in a Bottle truth set, so precision and recall were not measured.

### Future scope

- Use the same analysis on the full chr22 reads (around 30x coverage depth) to demonstrate increased sensitivity; the same approach can be applied to the whole genome using computational clusters or the cloud.
- Compare hard filtering with VQSR and with machine learning approaches.
- Automate the workflow using Nextflow (e.g. the nf-core/sarek pipeline) or Snakemake. This will make it fully reproducible.

## Data Availability

All data produced in the present work are contained in the manuscript

https://www.ebi.ac.uk/ena/browser/view/ERR3239334

## Notes

### Competing Interest Statement

The authors have declared no competing interest.

